# Coverage of primary and booster vaccination against COVID-19 by socioeconomic level: A nationwide cross-sectional registry study

**DOI:** 10.1101/2023.01.13.23284467

**Authors:** Bo T Hansen, Angela S Labberton, Prabhjot Kour, Kristian B Kraft

## Abstract

High and equitable COVID-19 vaccination coverage is important for pandemic control and prevention of health inequity. However, little is known about socioeconomic correlates of booster vaccination coverage. In this cross-sectional study of all Norwegian adults in the national vaccination program (N = 4,190,655), we use individual-level registry data to examine coverage by levels of household income and education of primary (**≥**2 doses) and booster (**≥**3 doses) vaccination against COVID-19. We stratify the analyses by age groups with different booster recommendations and report relative risk ratios (RR) for vaccination by 25 August 2022. In the 18-44 years group, individuals with highest vs. lowest education had 94% vs. 79% primary coverage (adjusted RR (adjRR) 1.15, 95%CI 1.14-1.15) and 67% vs. 38% booster coverage (adjRR 1.55, 95% CI 1.55-1.56), while individuals with highest vs. lowest income had 94% vs. 81% primary coverage (adjRR 1.10, 95%CI 1.10-1.10) and 60% vs. 43% booster coverage (adjRR 1.23, 95%CI 1.22-1.24). In the **≥**45 years group, individuals with highest vs. lowest education had 96% vs. 92% primary coverage (adjRR 1.02, 95%CI 1.02-1.02) and 88% vs. 80% booster coverage (adjRR 1.09, 95%CI 1.09-1.09), while individuals with highest vs. lowest income had 98% vs. 82% primary coverage (adjRR 1.16, 95%CI 1.16-1.16) and 92% vs. 64% booster coverage (adjRR 1.33, 95%CI 1.33-1.34). In conclusion, we document large socioeconomic inequalities in COVID-19 vaccination coverage, especially for booster vaccination, even though all vaccination was free-of-charge. The results highlight the need to tailor information and to target underserved groups for booster vaccination.

## Introduction

Mass vaccination is an essential strategy to control the COVID-19 pandemic because it may protect against infection, symptomatic disease, hospitalization and death associated with SARS-CoV-2 infection.^1,2^ However, vaccine effectiveness wanes over time and varies by virus variant.^3,4^ Additional vaccination, beyond the primary two doses, has proven effective in boosting the vaccine-induced immune response^5^ and in protecting against symptomatic disease, hospitalization and death.^4,6,7^ Many countries have thus added booster vaccination to their vaccination programs against COVID-19. High uptake of booster vaccination is important to prevent severe disease and death, as well as to prevent overburdening of the health care system. It may also reduce the need for imposing social restrictions. Many countries are already offering a second booster dose, and continued emergence of new variants and waning immunity by time since last vaccination is likely to make booster vaccination an important part of future COVID-19 vaccination programs.

Primary vaccination against COVID-19 has achieved high coverage in some European countries.^8^ Few studies have examined the relationship between primary COVID-19 vaccination and socioeconomic characteristics, especially with data covering the whole target population of a vaccination program. We have previously shown that program coverage of at least one vaccine dose is lower among some nationalities of immigrants to Norway, and their offspring, than among individuals born in Norway by Norwegian-born parents.^9^ A recent study of the general population in Sweden found that younger age, male sex, lower income and being born outside of Sweden were associated with lower uptake of at least one dose of COVID-19 vaccine.^10,11^ Registry-based studies from the UK also show social inequalities in uptake of at least one vaccine dose, including lower uptake among some ethnic groups and among individuals living in deprived areas.^12,13^ Self-reported survey data from the US also indicate inequalities in uptake of at least one dose of COVID-19 vaccine.^14,15^

Booster vaccination coverage is considerably lower than primary vaccination coverage,^8^ indicating more barriers to vaccination^16^ in the booster setting. A recent survey that addressed the intention for booster vaccination among adults that had completed primary vaccination in the UK found that lower socio-economic position was associated with uncertainty/unwillingness to receive a COVID-19 booster vaccine.^17^ Recent survey data from the US also indicate inequalities in self-reported booster vaccination uptake,^18^ and a population-based study of two large US metropolitan areas found inequalities in both primary and booster vaccination uptake by social vulnerability (measured by an area-level index), race/ethnicity and location.^19^

Ensuring good health for all and reducing inequalities, including in access to vaccines, are among the priorities of the World Health Organization (WHO).^20^ Addressing potential inequalities in COVID-19 vaccination uptake is important to inform interventions to minimize them, and thus the resulting inequalities in health. Low socio-economic status and ethnicity/country of birth are associated with more severe COVID-19 disease,^21-24^ and inequalities in primary and booster vaccine uptake may thus compound inequalities in health. In the present study, we utilize nationwide registries to examine socioeconomic correlates of primary and booster vaccination coverage of all adults in a national COVID-19 vaccination program.

## Methods

### Study setting

Norway has a relatively high total coverage of primary and booster vaccination against COVID-19 compared to most other countries.^8^ Vaccination included in the Corona Vaccination Program (CVP) is voluntary and free of charge. The main vaccines offered have been BNT162b2 mRNA (BioNTech/Pfizer) and mRNA-1273 (Moderna). Primary vaccination (two doses) has been recommended to those aged 65 and older since 21 December 2020, and to those aged 18 and older since 21 May 2021. Booster vaccination (third dose) has been recommended to those aged 65 and older since 5 October 2021, and to those aged 45 and older since 26 November 2021.^25^ Individuals aged 18-44 have also been offered booster vaccination in the CVP from 26 November 2021, but it is explicitly recommended only to those with underlying conditions that give an elevated risk for severe COVID-19 disease, while persons without medical risk conditions are offered booster vaccination for free if they wish to have it. For both vaccination settings throughout the study period, confirmed infection with SARS-CoV-2 has been considered equivalent to one vaccine dose in assessments of need for further doses. In the present study, vaccinations registered until 25 August 2022 were included, thus the follow-up time for primary and booster vaccination status was at least 15 and 9 months, respectively.

### Study design and population

In this population-based cross-sectional study, we used data from the Norwegian National Preparedness Register for COVID-19 (Beredt C19), which has been established for real-time surveillance and analysis of individual-level data relating to the pandemic. Beredt C19 contains data from different publicly owned data sources that cover the entire Norwegian population, including national health and administrative registries. Data from the various registries can be linked via the unique personal identification number given to each resident in Norway at birth or immigration. For this study, we linked individual-level data from the National Population Registry, the Norwegian Immunisation Register (SYSVAK), Statistics Norway (SSB), the Norwegian surveillance system for communicable diseases (MSIS), and a Beredt C19-specific variable of medical risk group classification generated by data from the Norwegian Patient Registry and the Norwegian Registry for Primary Health Care. We extracted data for the whole adult population of Norway (age 18 or older).

### Outcome variables

The outcome variables are having received at least two doses of vaccine against COVID-19, and having received at least three doses of vaccine against COVID-19, as registered in the Immunisation Register by 25 August 2022. Registration of vaccination in the Immunisation Register is mandated by law and considered complete for vaccination occurring in Norway. We do not distinguish between different types of COVID-19 vaccine. The CVP has almost exclusively offered COVID-19 vaccines that have a general recommendation of two doses for primary vaccination.^25^ Hence, at the population level, having received at least two vaccine doses equates to having completed primary vaccination, and having received at least three doses equates to having received booster vaccination.

### Predictor variables

Data on educational level and income was obtained from Statistics Norway and refers to the years 2019 and 2018, respectively, which is the most recent socioeconomic data available in Beredt C19 because there is a lag in registration of this data. The data on education refers to the highest attained education for each individual and was ordinally categorized into the following levels: Below upper secondary, Upper secondary, University short or University long. The two latter categories refer to undergraduate (up to four years) and postgraduate studies (more than four years). Individuals aged 18-25 years were allocated to a separate category because most of them would still be in education and may not have reached their highest educational level. The income data refers to total household income from all sources adjusted for household size and composition,^26^ categorized into deciles. Missing education or income data was coded as a separate category, thus the sample size is the same for all analyses of the same outcome.

Covariate data on age, sex and residency area was obtained from the National Population Registry. We categorized age into the groups 18-29, 30-39, 40-44, 45-49, 50-59, 60-69, 70-79, >=80. We categorized the residency covariate into living/not living in the capital area because Oslo has had the highest rates of notified infections throughout the pandemic,^27^ and urban living could be a factor in the spread of COVID-19.^28,29^ Covariate data on immigrant status was obtained from Statistics Norway. It has three categories: (i) Non-immigrant (i.e. have at least one Norwegian-born parent); (ii) Immigrant (i.e. foreign-born to two foreign-born parents); (iii) Norwegian-born to two foreign-born parents. When referring to the latter two groups collectively, we use the term “individuals with immigrant background” in the present paper.

Medical risk group is a Beredt C19-specific dichotomous covariate indicating whether an individual has one or more of a set of 14 diagnoses/health conditions identified as conveying a higher risk of hospitalization for COVID-19.^30^ Individuals with these conditions were prioritized for relatively early COVID-19 vaccination. In the CVP, confirmed SARS-CoV-2 infection has been considered as an immunological event equivalent to having had one dose of COVID-19 vaccine. Hence, in the analyses of the primary vaccination setting, we included a covariate for having had at least one confirmed SARS-CoV-2 infection before the CVP onset of second dose vaccination (yes/no), which was set to two months after the relevant age group became eligible for the first dose. Similarly, in the booster vaccination setting, we included a covariate for having had at least one confirmed SARS-CoV-2 infection before CVP onset of booster vaccination (yes/no). Individual data on confirmed SARS-CoV-2 infections were retrieved from the Norwegian surveillance system for communicable diseases.

### Statistics

We assessed associations between the socioeconomic variables and COVID-19 vaccination by modified Poisson regression with robust error variance, and present relative risks for being vaccinated versus not being vaccinated with 95% confidence intervals (95% CI). Primary and booster vaccination were analyzed as separate outcomes. We analyzed the age groups 18-44 years and >=45 years separately because they received different recommendations for booster vaccination (as described above). For each outcome, we present univariate analyses for education and income, as well as multivariate analyses mutually adjusted for all predictor variables. We also present sensitivity analyses of adjusted models that do not include variables indicating prior SARS-CoV-2 infection. Tests for trend were performed by the Jonckheere-Terpstra test. All tests were two-sided. P-values less than 0.05 were considered statistically significant. Data management and statistical computing was performed with Stata 17.0.

## Results

The study population comprised all persons aged 18 years or older who were resident in Norway per 1 January 2022 (N = 4,190,655), of which 1,853,123 were in the 18-44 years age group and 2,337,532 were in the >=45 years age group. The distribution of individuals by educational and income levels for each covariate is shown in Table 1.

**Table 1.** Education and income level in the adult population in Norway, by demographic and health-related variables. ^a^ Row percentage ^b^ Immigrant, or Norwegian-born with foreign-born parents ^c^ Registered diagnosis indicating elevated risk for hospitalization for COVID-19 ^d^ SARS-CoV-2 infection confirmed by laboratory ^e^ Highest attained education level ^f^ Individuals aged 18-25 years were categorized separately because many are undergoing education ^g^ Household income

|  | **N (% of total)** | **Mean age (years)** | **Male, N (%)<sup>a</sup> yes** | **Oslo residency, N (%)<sup>a</sup> yes** | **Immigrant background<sup>b</sup>, N (%)<sup>a</sup> yes** | **Medical risk group<sup>c</sup>, N (%)<sup>a</sup> yes** | **SARS-CoV-2<sup>d</sup> before 2<sup>nd</sup> dose recommendation, N (%)<sup>a</sup> yes** | **SARS-CoV-2<sup>d</sup> before 3<sup>rd</sup> dose recommendation, N (%)<sup>a</sup> yes** |
| --- | --- | --- | --- | --- | --- | --- | --- | --- |
| **Education level<sup>e</sup>** |  |  |  |  |  |  |  |  |
| <Upper secondary | 245597 (13) | 34 | 143846 (59) | 30799 (13) | 81884 (33) | 27720 (11) | 12651 (5) | 73635 (30) |
| Upper secondary | 399667 (22) | 35 | 246741 (62) | 37334 (9) | 67968 (17) | 38407 (10) | 13354 (3) | 123345 (31) |
| University short | 408051 (22) | 35 | 156539 (38) | 80897 (20) | 69592 (17) | 34794 (9) | 14001 (3) | 132312 (32) |
| University long | 198337 (11) | 36 | 88658 (45) | 59724 (30) | 52512 (26) | 13354 (7) | 5737 (3) | 61264 (31) |
| Unknown | 84583 (5) | 35 | 47052 (56) | 18367 (22) | 81442 (96) | 3758 (4) | 5563 (7) | 29885 (35) |
| 18-25 years<sup>f</sup> | 516888 (28) | 22 | 267297 (52) | 74859 (14) | 85090 (16) | 38152 (7) | 31355 (6) | 168724 (33) |
| **Income decile<sup>g</sup>** |  |  |  |  |  |  |  |  |
| 1 | 182514 (10) | 28 | 94282 (52) | 46254 (25) | 60212 (33) | 11760 (6) | 10436 (6) | 54561 (30) |
| 2 | 182514 (10) | 31 | 88829 (49) | 34524 (19) | 76864 (42) | 15842 (9) | 12154 (7) | 62737 (34) |
| 3 | 182512 (10) | 31 | 90584 (50) | 27938 (15) | 59808 (33) | 18197 (10) | 9402 (5) | 59219 (32) |
| 4 | 182515 (10) | 32 | 91035 (50) | 23463 (13) | 49399 (27) | 17100 (9) | 8009 (4) | 61387 (34) |
| 5 | 182512 (10) | 32 | 91819 (50) | 20915 (11) | 39171 (21) | 16965 (9) | 7263 (4) | 59923 (33) |
| 6 | 182514 (10) | 32 | 92518 (51) | 20535 (11) | 32012 (18) | 16393 (9) | 6552 (4) | 58594 (32) |
| 7 | 182513 (10) | 32 | 93762 (51) | 22530 (12) | 27473 (15) | 15986 (9) | 6614 (4) | 56837 (31) |
| 8 | 182515 (10) | 32 | 96394 (53) | 26423 (14) | 24705 (14) | 15382 (8) | 6345 (3) | 55923 (31) |
| 9 | 182511 (10) | 32 | 98307 (54) | 32107 (18) | 23118 (13) | 14339 (8) | 6472 (4) | 55131 (30) |
| 10 | 182513 (10) | 30 | 98400 (54) | 39826 (22) | 20264 (11) | 13560 (7) | 7812 (4) | 56014 (31) |
| Unknown | 27990 (2) | 32 | 14203 (51) | 7465 (27) | 25462 (91) | 661 (2) | 1602 (6) | 8839 (32) |
| **Total** | **1853123 (100)** | **31** | **950133 (51)** | **301980 (16)** | **438488 (24)** | **156185 (8)** | **82661 (4)** | **589165 (32)** |

**Table 1. Education and income level in the adult population in Norway, by demographic and health-related variables**
| Education level<sup>e</sup> |  |  |  |  |  |  |  |  |
| --- | --- | --- | --- | --- | --- | --- | --- | --- |
| <Upper secondary | 479142 (20) | 65 | 222617 (46) | 39562 (8) | 65714 (14) | 186645 (39) | 8456 (2) | 53277 (11) |
| Upper secondary | 1051332 (45) | 63 | 550630 (52) | 76170 (7) | 87579 (8) | 340327 (32) | 14606 (1) | 118347 (11) |
| University short | 548744 (23) | 60 | 227719 (41) | 70169 (13) | 58932 (11) | 135619 (25) | 9269 (2) | 80833 (15) |
| University long | 213014 (9) | 59 | 124732 (59) | 46664 (22) | 36411 (17) | 45791 (21) | 3731 (2) | 35585 (17) |
| Unknown | 45300 (2) | 57 | 24708 (55) | 9017 (20) | 37538 (83) | 9998 (22) | 2259 (5) | 10117 (22) |
| Income decile<sup>g</sup> |  |  |  |  |  |  |  |  |
| 1 | 233106 (10) | 64 | 101391 (43) | 33449 (14) | 74996 (32) | 77939 (33) | 6227 (3) | 33773 (14) |
| 2 | 233106 (10) | 66 | 102917 (44) | 22655 (10) | 37232 (16) | 90034 (39) | 3975 (2) | 28089 (12) |
| 3 | 233106 (10) | 65 | 110826 (48) | 19659 (8) | 30051 (13) | 85977 (37) | 3430 (1) | 27961 (12) |
| 4 | 233104 (10) | 64 | 115575 (50) | 18866 (8) | 26384 (11) | 80231 (34) | 3385 (1) | 28473 (12) |
| 5 | 233106 (10) | 63 | 117304 (50) | 18945 (8) | 23425 (10) | 74722 (32) | 3449 (1) | 28911 (12) |
| 6 | 233106 (10) | 62 | 117577 (50) | 19994 (9) | 21534 (9) | 69887 (30) | 3500 (2) | 29652 (13) |
| 7 | 233104 (10) | 61 | 118604 (51) | 21240 (9) | 19164 (8) | 65950 (28) | 3430 (1) | 29150 (13) |
| 8 | 233107 (10) | 61 | 119370 (51) | 23104 (10) | 16961 (7) | 61949 (27) | 3522 (2) | 29140 (13) |
| 9 | 233105 (10) | 60 | 120313 (52) | 26101 (11) | 15967 (7) | 57870 (25) | 3278 (1) | 29313 (13) |
| 10 | 233104 (10) | 60 | 122803 (53) | 36315 (16) | 15213 (7) | 53068 (23) | 3869 (2) | 32283 (14) |
| Unknown | 6478 (0) | 54 | 3726 (58) | 1254 (19) | 5247 (81) | 753 (12) | 256 (4) | 1414 (22) |
| Total | 2337532 (100) | 62 | 1150406 (49) | 241582 (10) | 286174 (12) | 718380 (31) | 38321 (2) | 298159 (13) |

Among individuals aged 18-44 years, 1,633,499 (88%) had completed primary vaccination, and 959,958 (52%) had completed booster vaccination by 25 August 2022 (Table 2). The corresponding numbers among individuals aged >=45 years were 2,203,115 (94%) and 1,978,236 (85%) (Table 3).

**Table 2:**
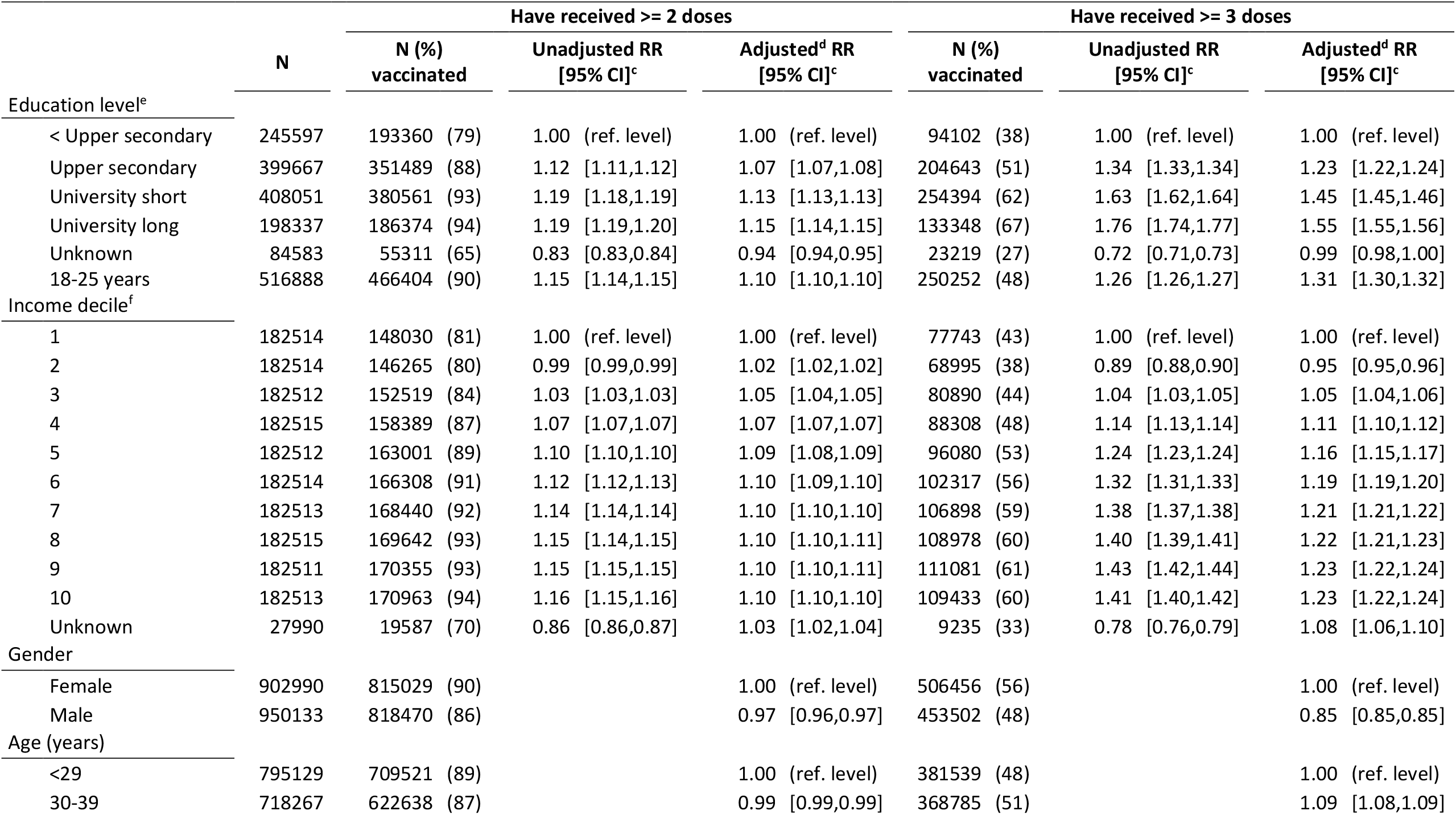
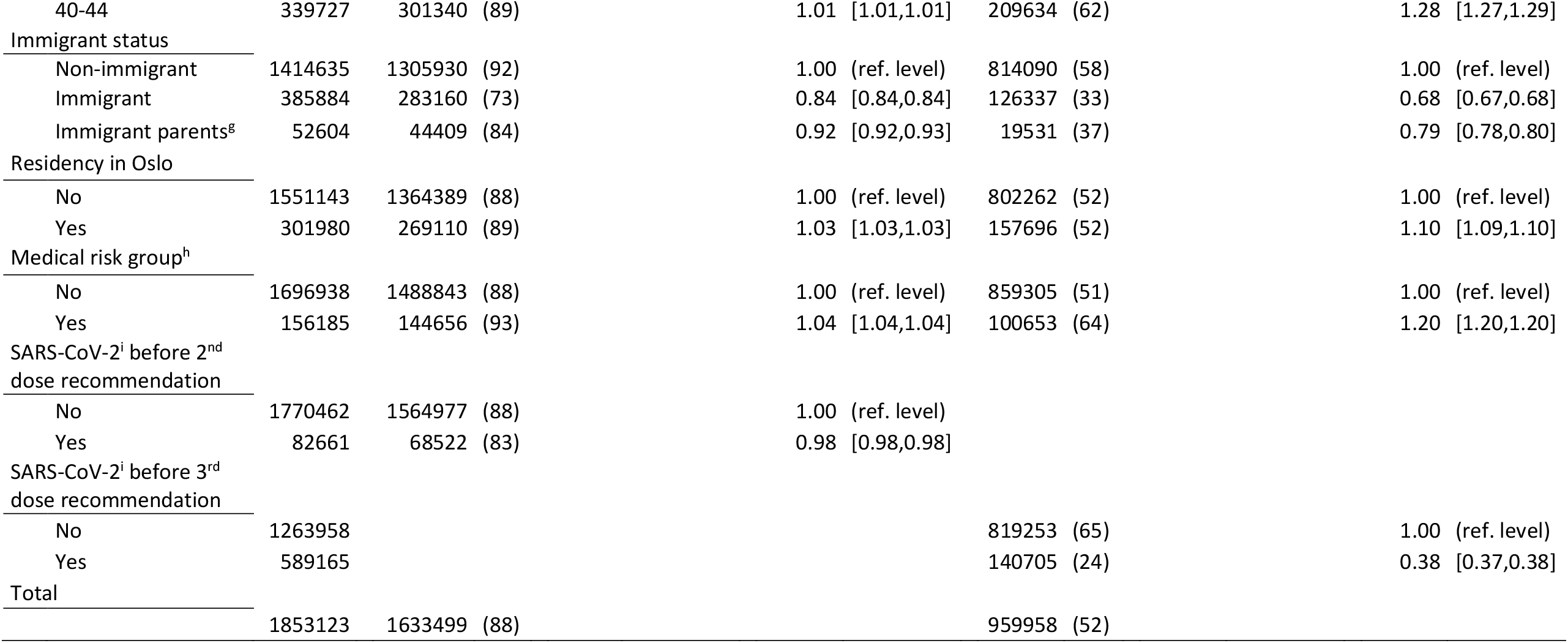
Regression analyses^a^ for completion of primary vaccination (>=2 doses) and booster vaccination (>=3 doses) against COVID-19 by sociodemographic and health characteristics among individuals 18-44 years of age^b^. ^a^ Modified Poisson regression ^b^ All COVID-19 vaccination by 25 August 2022 among all Norwegian residents in the age bracket ^c^ RR = relative risk ratio, CI = confidence interval ^d^ Mutually adjusted for all tabulated variables ^e^ Highest attained education level ^f^ Household income ^g^ Norwegian-born with immigrant parents ^h^ Registered diagnosis indicating elevated risk of hospitalization for COVID-19 ^i^ SARS-CoV-2 infection confirmed by laboratory

**Table 3:**
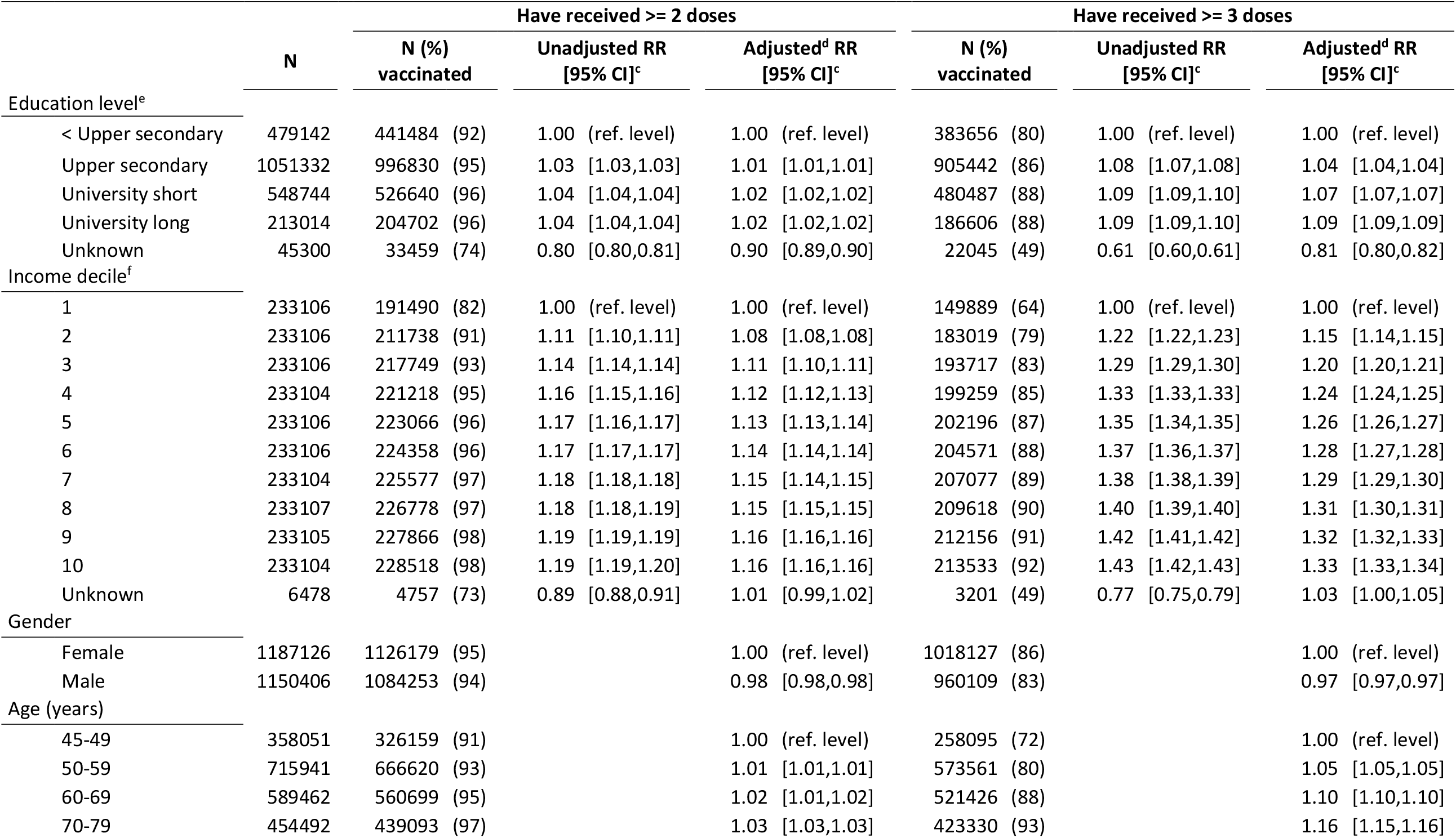
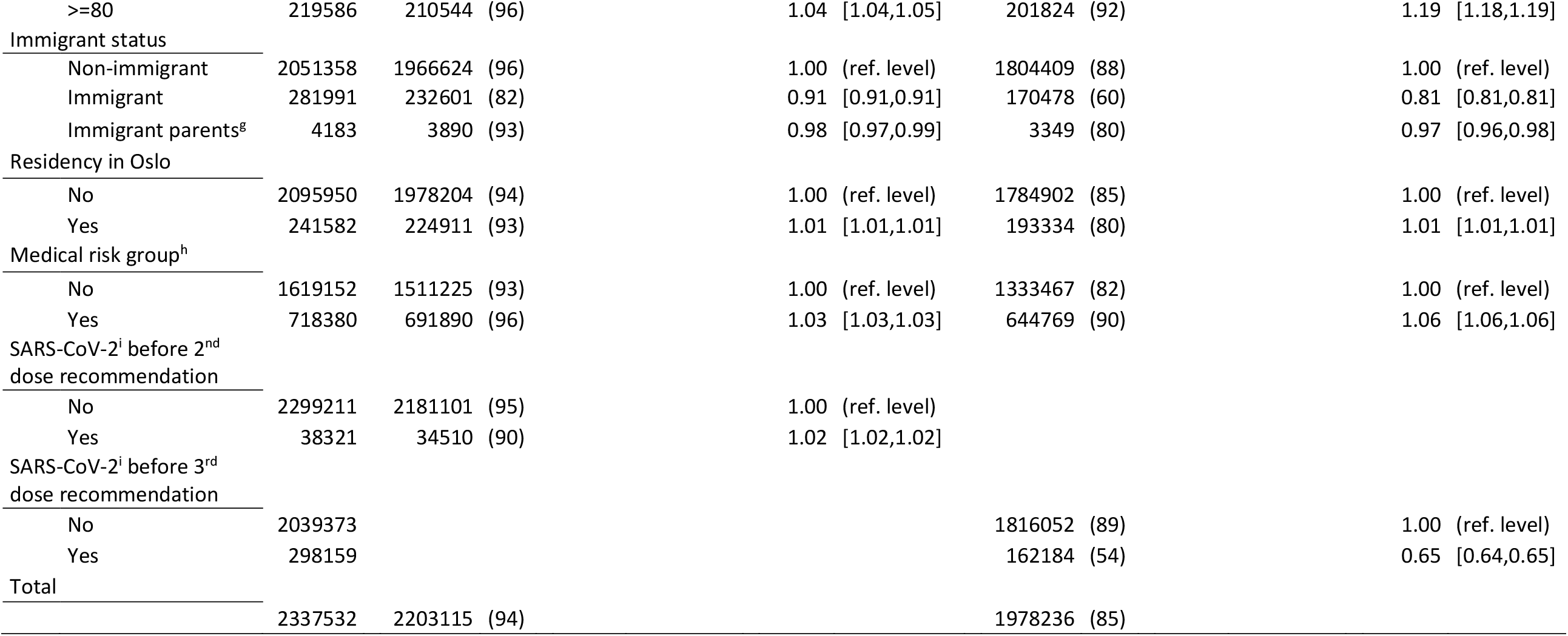
Regression analyses^a^ for completion of primary vaccination (>=2 doses) and booster vaccination (>=3 doses) against COVID-19 by sociodemographic and health characteristics among individuals >=45 years of age^b^. ^a^ Modified Poisson regression ^b^ All COVID-19 vaccination by 25 August 2022 among all Norwegian residents in the age bracket ^c^ RR = relative risk ratio, CI = confidence interval ^d^ Mutually adjusted for all tabulated variables ^e^ Highest attained education level ^f^ Household income ^g^ Norwegian-born with immigrant parents ^h^ Registered diagnosis indicating elevated risk of hospitalization for COVID-19 ^i^ SARS-CoV-2 infection confirmed by laboratory

### Coverage by education level

In both vaccination settings and in both age groups, vaccination coverage was higher with increasing education level (all Ps for trend among individuals with known educational level <0.0001). The lowest coverage was observed among individuals with unknown education level. Unadjusted and adjusted models showed similar differences and trends in RRs by education level, although the effects were somewhat smaller in adjusted models (Tables 2, 3). Adjustment had greatest effect on the RR estimates by education level in the booster vaccination setting, and in the 18-44 years age group. It also affected the unknown education level disproportionately. Absolute and relative differences between education levels were larger in the booster than in the primary vaccination setting (Figure 1). For both vaccination settings, the differences in coverage by education level were smaller in the >=45 years age group than in the 18-44 years age group (Figure 1). In the 18-44 years age group (Table 2, Figure 1A, B), the primary vaccination coverage was 94% among individuals with long university education, and 79% among those with less than upper secondary education. The resulting adjusted RR (adjRR) for being vaccinated among individuals with the highest versus lowest educational attainment was 1.15 (95% CI: 1.14 to 1.15). For booster vaccination, the corresponding coverages were 67% and 38%, which yielded an adjRR of 1.55 (1.55 to 1.56). In the >=45 years age group (Table 3, Figure 1C, D), the primary vaccination coverage was 96% among individuals with long university education, and 92% among those with less than upper secondary education. The resulting adjRR for being vaccinated among individuals with the highest versus lowest educational attainment was 1.02 (1.02 to 1.02). For booster vaccination, the corresponding coverages were 88% and 80%, which yielded an adjRR of 1.09 (1.09 to 1.09).

**Figure 1.**
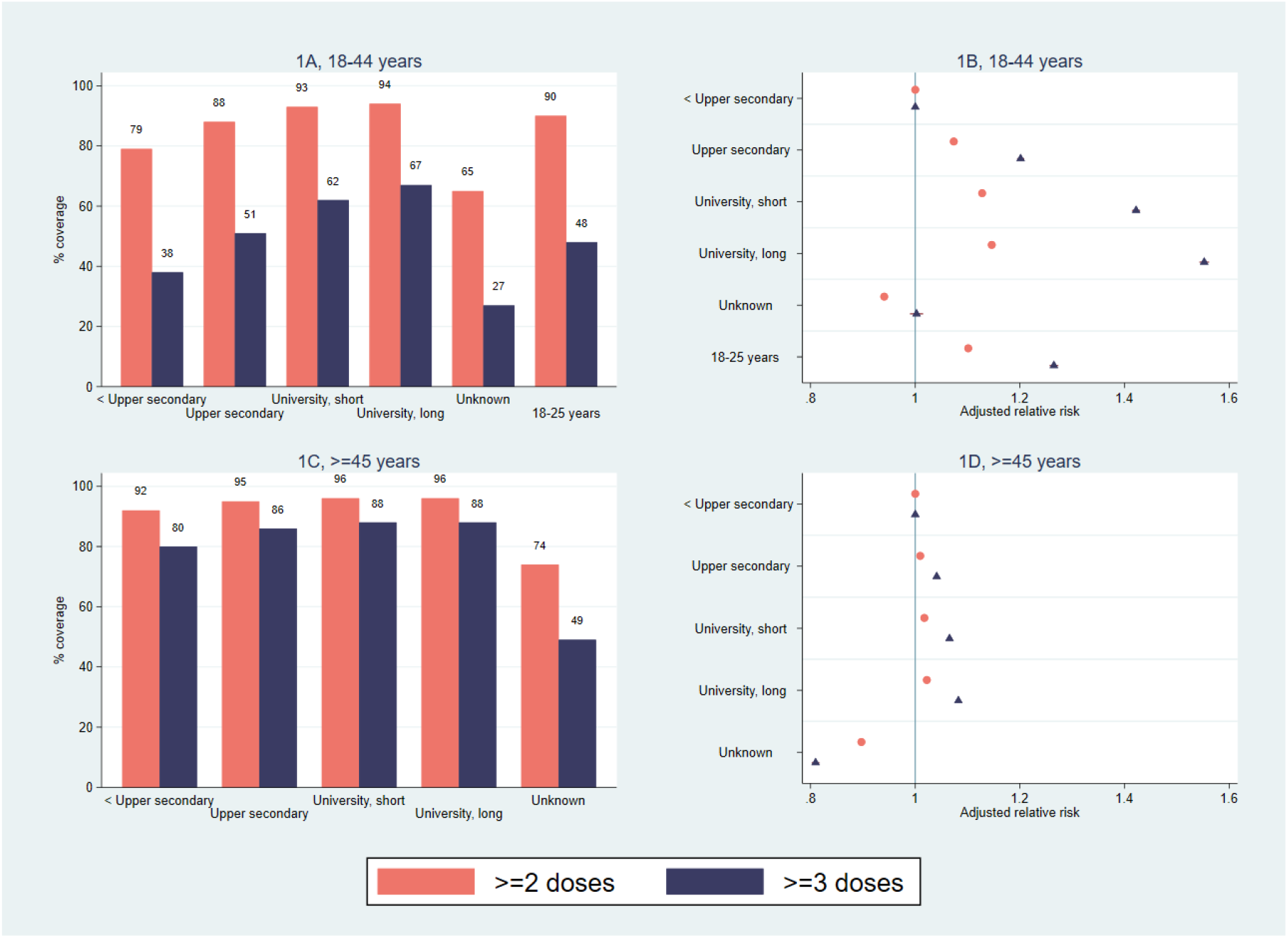
COVID-19 vaccination coverage for >=2 doses (red bars/circles) and >=3 doses (blue bars/triangles), by highest attained education level. A) Vaccination coverage (%) for individuals aged 18-44 years. B) Adjusted relative risks for vaccination with associated 95% confidence intervals among individuals aged 18-44 years. C) Vaccination coverage (%) for individuals aged >=45 years. D) Adjusted relative risks for vaccination with associated 95% confidence intervals among individuals aged >=45 years

### Coverage by income level

Similar overall patterns were observed by income level as by educational level (Figures 1, 2). In both vaccination settings and in both age groups, vaccination coverage was generally higher with increasing income level (all Ps for trend among individuals with known educational level <0.0001), although individuals in the second income decile had a slightly lower coverage than individuals in the first income decile. The lowest coverage was observed among individuals with unknown income level. Adjustment tended to attenuate the effects observed in unadjusted models, but adjusted models showed a consistently higher effect by increasing income level (Tables 2, 3). Adjustment had greatest effect on the RR estimates by income level in the booster vaccination setting, and in the 18-44 years age group. It also affected the unknown income level disproportionately. Absolute and relative differences between income levels were larger in the booster than in the primary vaccination setting (Figure 2). For both vaccination settings, the absolute and relative differences by income levels were somewhat larger in the >=45 years age group than in the 18-44 years age group (Figure 2). In the 18-44 years age group (Table 2, Figure 2A, B), the primary vaccination coverage was 94% among individuals in the 10^th^ (i.e. highest) income decile, and 81% among those in the 1^st^ (i.e. lowest) income decile. The resulting adjRR for being vaccinated among individuals with the highest versus lowest income was 1.10 (1.10 to 1.10). For booster vaccination, the corresponding coverages were 60% and 43%, which yielded an adjRR of 1.23 (1.22 to 1.24). In the >=45 years age group (Table 3, Figure 2C, D), the primary vaccination coverage was 98% among individuals in the 10^th^ income decile, and 82% among those in the 1^st^ income decile. The resulting adjRR for being vaccinated among individuals with the highest versus lowest income level was 1.16 (1.16 to 1.16). For booster vaccination, the corresponding coverages were 92% and 64%, which yielded an adjRR of 1.33 (1.33 to 1.34).

**Figure 2.**
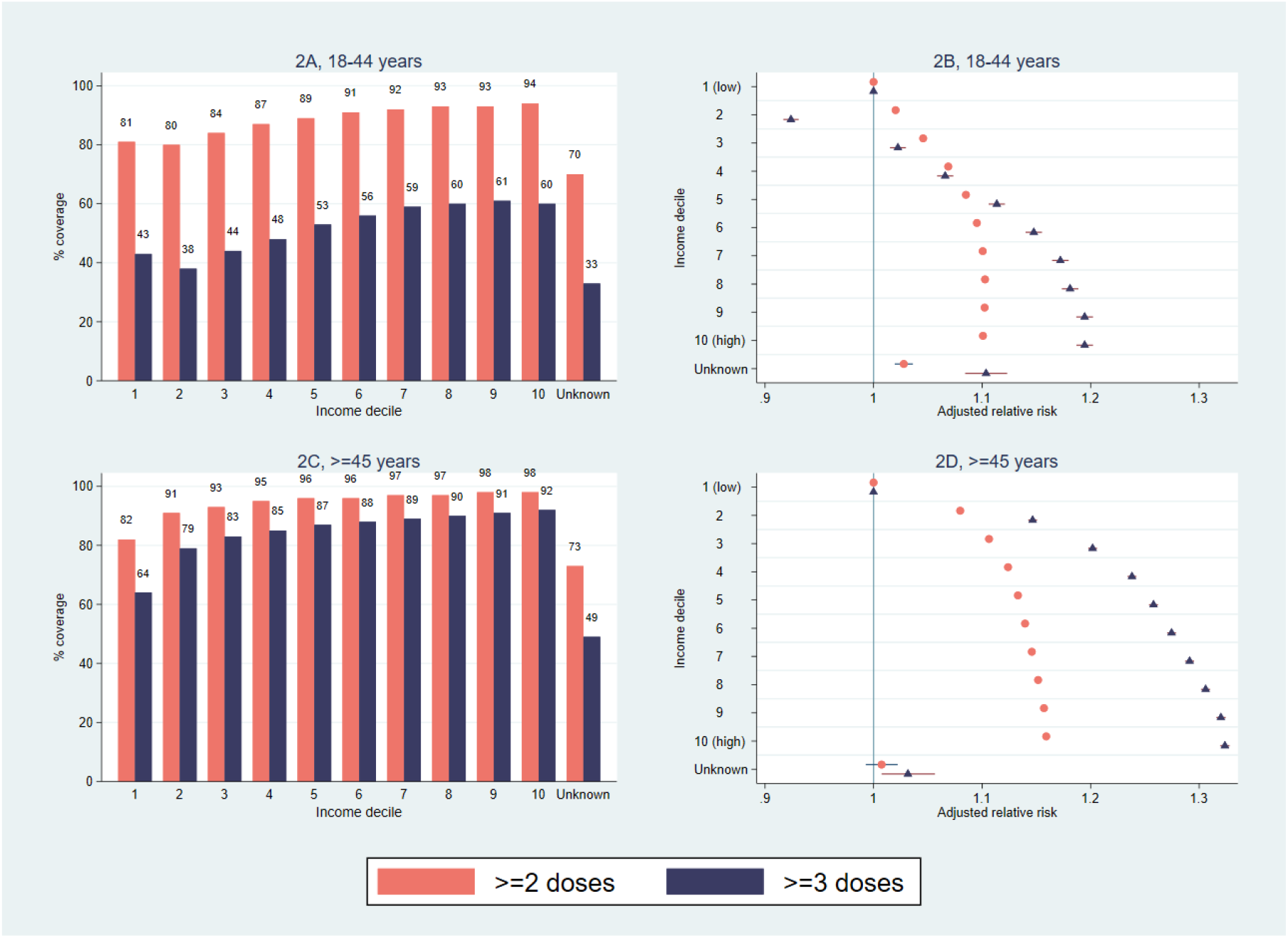
COVID-19 vaccination coverage for >=2 doses (red bars/circles) and >=3 doses (blue bars/triangles), by household income level. A) Vaccination coverage (%) for individuals aged 18-44 years. B) Adjusted relative risks for vaccination with associated 95% confidence intervals among individuals aged 18-44 years. C) Vaccination coverage (%) for individuals aged >=45 years. D) Adjusted relative risks for vaccination with associated 95% confidence intervals among individuals aged >=45 years

### Covariate effects

The covariates had overall similar effects in the 18-44 and >=45 years age groups. Relatively small differences between levels of each covariate were observed in the primary vaccination setting, except for by immigrant status, where individuals with immigrant background had lower coverage than non-immigrants. However, in the booster vaccination setting, differences exceeding 10 percentage point were observed by levels of age, immigrant status, medical risk group and past SARS-CoV-2 infection (Tables 2, 3). Booster vaccination coverage increased by age, was lower among individuals with immigrant background, was higher among individuals with elevated risk of hospitalization for COVID-19, and among individuals who had not had COVID-19 before onset of vaccination. Adjusted models that did not include variables indicating SARS-CoV-2 infection before onset of vaccination gave very similar results as the fully adjusted models in both age strata and in both vaccination settings (Supplementary table 1).

**Supplementary table 1:**
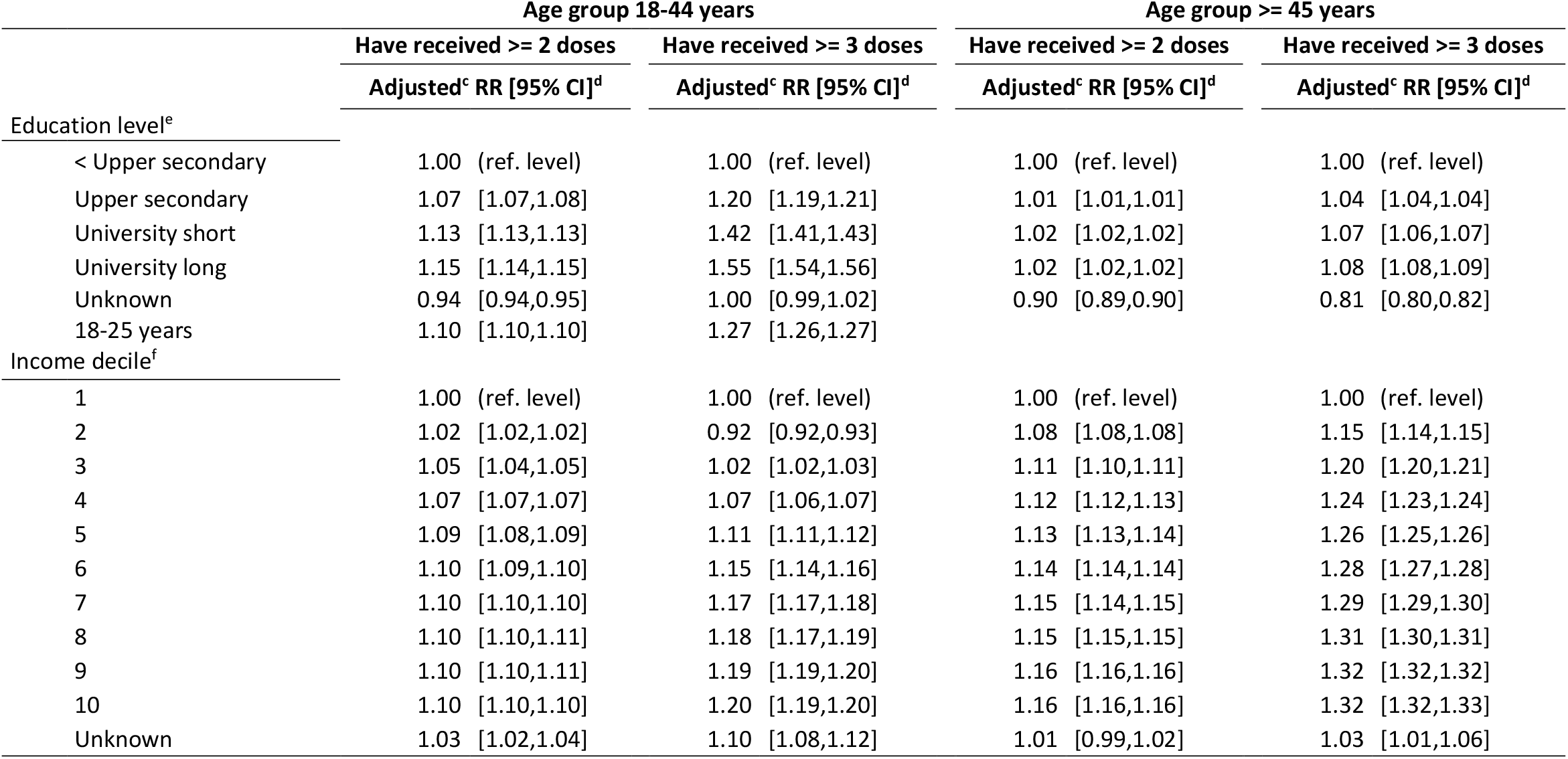
Regression analyses^a^ for completion of primary vaccination (>=2 doses) and booster vaccination (>=3 doses) against COVID-19^b^ by socioeconomic level. *The estimates are not adjusted by confirmed SARS-CoV-2 infection prior to onset of vaccination*. ^a^ Modified Poisson regression, stratified by age group (18-44 years, >=45 years) ^b^ All COVID-19 vaccination by 25 August 2022 among all Norwegian residents aged 18 and older ^c^ Estimates adjusted by gender, age, immigrant status, residency in Oslo, medical risk group, and education (for income) and income (for education). See Tables 2 and 3 in main paper for details ^d^ RR = relative risk ratio, CI = confidence interval ^e^ Highest attained education level ^f^ Household income

|  | Age group 18-44 years |  | Age group >= 45 years |  |
| --- | --- | --- | --- | --- |
|  | Have received >= 2 doses | Have received >= 3 doses | Have received >= 2 doses | Have received >= 3 doses |
|  | Adjusted<sup>c</sup> RR [95% CI]<sup>d</sup> | Adjusted<sup>c</sup> RR [95% CI]<sup>d</sup> | Adjusted<sup>c</sup> RR [95% CI]<sup>d</sup> | Adjusted<sup>c</sup> RR [95% CI]<sup>d</sup> |
| Education level<sup>e</sup> |  |  |  |  |
| < Upper secondary | 1.00 (ref. level) | 1.00 (ref. level) | 1.00 (ref. level) | 1.00 (ref. level) |
| Upper secondary | 1.07 [1.07,1.08] | 1.20 [1.19,1.21] | 1.01 [1.01,1.01] | 1.04 [1.04,1.04] |
| University short | 1.13 [1.13,1.13] | 1.42 [1.41,1.43] | 1.02 [1.02,1.02] | 1.07 [1.06,1.07] |
| University long | 1.15 [1.14,1.15] | 1.55 [1.54,1.56] | 1.02 [1.02,1.02] | 1.08 [1.08,1.09] |
| Unknown | 0.94 [0.94,0.95] | 1.00 [0.99,1.02] | 0.90 [0.89,0.90] | 0.81 [0.80,0.82] |
| 18-25 years | 1.10 [1.10,1.10] | 1.27 [1.26,1.27] |  |  |
| Income decile<sup>f</sup> |  |  |  |  |
| 1 | 1.00 (ref. level) | 1.00 (ref. level) | 1.00 (ref. level) | 1.00 (ref. level) |
| 2 | 1.02 [1.02,1.02] | 0.92 [0.92,0.93] | 1.08 [1.08,1.08] | 1.15 [1.14,1.15] |
| 3 | 1.05 [1.04,1.05] | 1.02 [1.02,1.03] | 1.11 [1.10,1.11] | 1.20 [1.20,1.21] |
| 4 | 1.07 [1.07,1.07] | 1.07 [1.06,1.07] | 1.12 [1.12,1.13] | 1.24 [1.23,1.24] |
| 5 | 1.09 [1.08,1.09] | 1.11 [1.11,1.12] | 1.13 [1.13,1.14] | 1.26 [1.25,1.26] |
| 6 | 1.10 [1.09,1.10] | 1.15 [1.14,1.16] | 1.14 [1.14,1.14] | 1.28 [1.27,1.28] |
| 7 | 1.10 [1.10,1.10] | 1.17 [1.17,1.18] | 1.15 [1.14,1.15] | 1.29 [1.29,1.30] |
| 8 | 1.10 [1.10,1.11] | 1.18 [1.17,1.19] | 1.15 [1.15,1.15] | 1.31 [1.30,1.31] |
| 9 | 1.10 [1.10,1.11] | 1.19 [1.19,1.20] | 1.16 [1.16,1.16] | 1.32 [1.32,1.32] |
| 10 | 1.10 [1.10,1.10] | 1.20 [1.19,1.20] | 1.16 [1.16,1.16] | 1.32 [1.32,1.33] |
| Unknown | 1.03 [1.02,1.04] | 1.10 [1.08,1.12] | 1.01 [0.99,1.02] | 1.03 [1.01,1.06] |

## Discussion

We have shown that there are socioeconomic inequalities in COVID-19 vaccination coverage among adults eligible for free vaccination in a nationwide vaccination program. There was a distinct trend of decreasing coverage by decreasing income and educational level. Moreover, the inequalities observed were larger for booster vaccination than for completion of primary vaccination. The relative differences were still evident after adjustment for a range of potentially confounding factors. We observed similar overall patterns by socioeconomic levels in the 18-44 and the >=45 years age groups. However, the coverage was higher, and the coverage differences by education level were smaller in the older age group. Large differences in coverage were also observed by age and by immigrant status, with a particularly low booster vaccination coverage among younger age groups and among immigrants.

To our knowledge, this is the first study to examine socioeconomic differences of primary and booster vaccination coverage with individual vaccination, socioeconomic, demographic and health data for an entire adult population in a national vaccination program. Surveys from the UK and US have reported social inequalities in the intention to receive^17^ and in self-reported COVID-19 booster vaccination.^18^ Here we document social inequalities with real-world data. Although direct comparisons are hampered by differences in context, populations, available data and analytic approach, our results generally coincide with the findings from a population-based study from two metropolitan areas in the US, including the observation that inequalities were larger for booster than for primary vaccination.^19^ Unique features of our study include the nationwide population and the use of individual data on educational level and household income.

Factors that may influence decisions regarding vaccination include trust in the effectiveness/safety of vaccines and/or in the system that delivers them, perception of disease risk, structural and psychological factors related to access and pragmatics, proneness to extensive information searching, and willingness to protect others.^31^ Our study does not address the root causes of coverage inequalities, and we thus do not know the relative importance of these potential determinants for the coverage differences we observe. However, we suggest that it might be associated to the lower priority given to booster vaccination, which is reflected by the overall lower coverage observed in this vaccination setting. Booster vaccination may have been perceived as less urgent because its campaign has been less intensive. Moreover, booster vaccination has been recommended and offered to a smaller part of the population. A lower booster vaccination coverage among individuals 18-44 years is expected, since they were offered the booster in the program without the explicit recommendation for vaccination conveyed to those aged >=45 years. A less intensive campaign with less social nudging may result in coverage inequalities because it may not be as successful in reaching disadvantaged parts of the population with information about the necessity for booster vaccination, how it can be achieved, and the fact that it is free of charge. A lower perceived urgency for booster vaccination may also induce coverage inequalities because other activities may take priority and displace vaccination, especially among those with limited resources. In many locations, booster vaccination was somewhat less accessible because there have been fewer administration sites and shorter opening hours after most of the population had completed primary vaccination. Lower accessibility might induce socioeconomic inequalities, for instance by increasing travel time and costs. Increased socioeconomic inequalities in health may thus be an unfortunate side effect of the lower priority given to booster vaccination. Studies addressing the causes of socioeconomic inequalities of COVID-19 booster vaccination uptake, and the adequacy of interventions that may diminish their impact, are urgently needed to avoid perpetuating inequalities in health.

All data used in this study were extracted from nationwide registries, which ensures objective and precise data routinely collected in a standardized manner for the entire population. There is thus no selection or response biases associated with the data presented here. Individual-level data was used in all analyses, which precludes ecological fallacy and increases the precision of the inference. Moreover, this study had a large sample size and thus high power to detect differences in COVID-19 primary and booster vaccination coverage.

Although we present analyses that were adjusted for a range of characteristics that might be associated with socioeconomic level and COVID-19 vaccine uptake, some residual confounding by characteristics unaccounted for in our data is likely. We adjusted for past laboratory-confirmed infection, which is a strength of this study, but did not have data on self- or undiagnosed COVID-19. However, adjustment for confirmed infection before onset of vaccination, which included nearly 900,000 infected individuals in the booster vaccination analyses, had very little impact on the education and income effect estimates, indicating little confounding by past infection on the observed coverage differences by socioeconomic level. Another limitation is that the socioeconomic data was from 2018/2019 while the coverage data was from 2021/2022. A small proportion of individuals in the target population also had missing socioeconomic data, of which many had immigrant background. Further limitations relate to potential misclassification of vaccination status. First, we did not have access to data of COVID-19 vaccination that may have occurred outside Norway and has not been registered in Norway at the vaccinee’s initiative. However, the extent of vaccination abroad among immigrants to Norway appears to be very limited.^32^ Second, we were not able to identify severely immunocompromised individuals in our dataset, who are recommended three doses to complete primary vaccination. However, they only account for a small part of the population,^27^ hence the impact of this bias is likely to be small. Moreover, our presentation of coverage rates by more than two and more than three doses may facilitate comparison with online vaccination trackers and other studies.

In conclusion, using individual-level data covering the whole adult population of Norway, we document large socioeconomic inequalities in free-of-charge COVID-19 booster vaccination coverage, in a country that has a relatively high general compliance to vaccine recommendations. Low booster coverage was observed among individuals with low income, low education, or who had immigrant background. Inequalities of smaller magnitude also persisted for completion of primary vaccination. Boosters improve public health and may remain important to curb COVID-19 in the coming years, hence research is needed to document why there are large socioeconomic inequalities for COVID-19 booster vaccination. Measures that increase booster vaccination coverage in general, and especially among the underrepresented groups identified here, could minimize inequalities in health. Increasing the accessibility of booster vaccination may reduce the direct and indirect costs for vaccinees, which could benefit individuals with low socioeconomic status.

## Data Availability

The data contains personal information and data access is only possible by ethics committee
approval and appropriate application to the public registries used in this study.

## Author contributions

**Bo T Hansen**: Conceptualization, Methodology, Validation, Writing – original draft, Writing – review and editing. **Angela S Labberton**: Methodology, Writing – review and editing. **Prabhjot Kour**: Methodology, Writing – review and editing. **Kristian B Kraft**: Methodology, Formal analysis, Data curation, Writing – review and editing

## Ethical approval

Ethical approval for the present study was obtained from the Regional Committee for Medical and Health Research Ethics, Oslo, Norway (REK sør-øst, reference number #198964). Requirement for consent was waived by the ethics committee.

## Data availability

The data contains personal information and data access is only possible by ethics committee approval and appropriate application to the public registries used in this study.

